# 6-Month Recovery after Mild Traumatic Brain Injury in Older Adults: A TRACK-GERI Study

**DOI:** 10.64898/2026.06.25.26356520

**Authors:** Yael Rosen-Lang, Ava M. Puccio, Esther L. Yuh, Domenico Lombardi, Katherine Kuang, Michele Diaz Nelson, W. John Boscardin, J. Russell Huie, Kristine Yaffe, David O. Okonkwo, Ramon Diaz Arrastia, Geoffrey T. Manley, Raquel C. Gardner, the Transforming Research and Clinical Knowledge in Traumatic Brain Injury (TRACK-TBI) investigators

## Abstract

**Importance:** Older adults (≥65 years old) are the largest and fastest growing population of traumatic brain injury (TBI) victims. However, real-world evidence of 6-month functional outcomes is limited.

**Objective:** To describe 6-month functional outcomes in older adults after mild TBI (mTBI; presenting Glasgow Coma Scale score 13-15), by age group and preinjury cognitive status. To evaluate the performance of the Glasgow Outcome Scale–Extended (GOSE) as a geriatric mTBI clinical trial endpoint.

**Design:** Prospective cohort study; single-center pilot enrollment February 2018-April 2019; two-center enrollment October 2020-March 2025.

**Setting:** Two U.S. level I trauma centers.

**Participants:** Older adults presenting within 72 hours of mTBI who received head CT, and their co-enrolled study-partner informants. Data curation and analysis were performed January 2023-November 2025.

**Exposure:** mTBI.

**Main outcome and measures:** GOSE was assessed at 2 weeks and 6 months post-injury and weighted using inverse probability weighting to mitigate attrition bias. Reliable change index was used to estimate the proportions of GOSE improvement or deterioration from 2 weeks to 6 months versus a 9-item Activities of Daily Living survey (ADL, Supplemental Methods) and Functional Activities Questionnaire (FAQ).

**Results:** Among 253 participants with mTBI, mean (SD) age was 77.3 (8.3) years, 129 (51%) were female, 68 (34%) had pre-injury mild cognitive impairment and 28 (15%) had pre-injury dementia (58 had unknown preinjury cognitive status). At 6 months, 14% died, 73% achieved home independence (GOSE 5+), and 17% achieved complete recovery (GOSE 8). Younger-old and persons without preinjury cognitive impairment had substantially better recovery (mortality 6-11%, home independence 84-86%, complete recovery 21-22%) versus oldest-old and those with pre-injury dementia (mortality 29-32%, home independence 23-52%, complete recovery 5-10%). GOSE detected the greatest proportion of reliable improvement or deterioration compared to ADL and FAQ, though low agreement reflects their complementary domains.

**Conclusions and Relevance:** Most younger and cognitively-unimpaired older adults achieved favorable functional outcomes, however, more vulnerable older adults may suffer chronic disability after mTBI. GOSE effectively captures meaningful change in this population, while ADL and FAQ provide important complementary information. Findings underscore the need for age-appropriate prognostication and more inclusive geriatric TBI research and clinical trial design.

**Trial Registration:** NCT07296783

**Key points:** *Question:* What are the functional outcomes of older adults 6 months after mild TBI, by age group and preinjury cognitive status?

*Findings:* Most older adults achieved home independence, though complete recovery was less common and mortality was notable. Younger-old and preinjury cognitively unimpaired individuals had substantially better recovery, with lower mortality and higher rates of home independence and complete recovery, compared to the oldest-old and those who had dementia preinjury.

*Meaning:* 6 months after mild TBI, the younger-old and cognitively-resilient have high potential for favorable outcome, while the older-old and cognitively-vulnerable are at high risk for mortality or chronic disability.

## Introduction

Global incidence of traumatic brain injury (TBI) among older adults, defined here as individuals age 65 years and older, is high and continues to rise despite falling incidence in younger age groups.1,2 We previously reported that 1 in 10 older adult Medicare beneficiaries had a TBI-related emergency department (ED) visit between 2000-2018.3 Older adults comprise an estimated 40% of TBI-related deaths in the U.S.4 and an estimated 38% of TBI-related ED visits in Europe,5 far exceeding their 18-22% proportion of the general U.S.6 and European populations.7 Although over 80% of TBIs in older adults are categorized as “mild” (e.g., presenting Glasgow Coma Scale [GCS] score 13-15),8,9 older adult TBI is associated with substantial chronic disability, institutionalization,^10^ dementia,^11^ and death.^12^,13

Yet, TBI outcomes in real-world older adults have been understudied. Large multi-center prospective cohort studies and clinical trials of TBI often exclude persons over age 6514. Other common exclusion criteria are pre-existing medical or neurological conditions that may confound outcomes, specifically dementia and mild cognitive impairment (MCI), whose prevalence increase with age.15 The European 65-center CENTER-TBI16 study and the U.S. 18-center TRACK-TBI17 study both excluded participants who had pre-existing neurological disorders such as dementia that could confound outcome assessments or who arrived to the ED >24 hours post-injury. These exclusions may differentially exclude older adults, many of whom have pre-existing conditions5,18,19 and may arrive later to the ED.20 Ultimately, the proportion of participating older adults was 28% in CENTER-TBI and 12% in TRACK-TBI, far below the expected proportion of TBI-related ED visits in older adults.4,5,21

Transforming Research And Clinical Knowledge in Geriatric Traumatic Brain Injury (TRACK-GERI) aimed to fill this knowledge gap and provide real-world evidence on TBI outcomes in older adults. This two-center prospective cohort study enrolled 270 adults age ≥65 years, regardless of pre-existing medical or neurological conditions, who presented to the ED within 72 hours of head trauma and received a head CT. In this analysis, we report the primary global functional outcome: the 6-month Glasgow Outcome Scale–Extended (GOSE), by age group and preinjury cognitive status. We further report on 6-month GOSE trajectories and performance as a geriatric TBI clinical trial endpoint. The over-arching aim of this analysis is to directly inform more precise and age-appropriate prognostication in real-world geriatric TBI and to facilitate the design of representative, adequately-powered clinical trials to improve outcomes in this under-studied and burdened population.

## Methods

### Study design and participants

The TRACK-GERI study enrolled 270 older adults who presented to the ED within 72 hours of head trauma and received a head CT, and 90 demographically-similar uninjured controls. Participants with TBI and controls were co-enrolled with a study partner informant. All enrolled (participants with TBI, controls, study partners) or their legally-authorized representatives completed informed consent. Additional inclusion criteria included being English or Chinese-speaking. Participant and study partner dyads were followed at 2-weeks and 3, 6, and 12 months post-injury with geriatric, functional, and neuropsychiatric outcome assessments performed by trained examiners. Since only 6% of TBI participants presented with GCS score <13 (n=15) or missing (n=2), then this analysis focused on those presenting with GCS score 13-15 (n=253) to facilitate more precise generalizability of our findings. Data from controls were used as a reference for 6-month normative change in outcomes. Ethical approval was obtained from the UCSF Institutional Review Board (#19-28925).

### Data types

#### Baseline characteristics

We collected participants’ demographics, medical history, injury and clinical characteristics on arrival to the ED (details in **Supplementary Methods**). We assessed pre-injury cognitive function using the informant-reported Clinical Dementia Rating (CDR) interview.22 CDR global score was used to categorize pre-injury cognitive status as cognitively unimpaired (CDR=0), mild cognitive impairment (MCI; CDR=0.5), or dementia (CDR > 0.5). Every CDR was reviewed and adjudicated by a behavioral neurologist (RCG) or geriatric nurse practitioner (MDN). We also collected study partners’ demographics and relationship and frequency of contact with the study participant.

#### Primary Outcome

6 month GOSE23 was our primary outcome. GOSE assesses functional recovery, ranging between 1 (dead) to 8 (complete recovery to pre-injury functional status), and is widely used as the primary outcome measure in TBI research and trials.16,24,25 It is assessed using a semi-structured interview in person or over the phone26 and may be completed by the participant or an informant, thus optimizing data collection. As in CENTER-TBI and most TRACK-TBI studies reporting on functional outcome, we used GOSE-all rather than GOSE-TBI27 given the challenges of discriminating TBI-related versus extracranial-injury-related deficits in this population. GOSE was assessed with both the participant and study partner if possible, and the lower score was used. A single universal GOSE threshold has not been established.28 To optimize statistical power and for outcome interpretability we report both the full GOSE score and the GOSE score dichotomized at independence at home (GOSE 5+ vs <5) at 2-weeks and 6-months. Every GOSE score was reviewed and adjudicated by the local research team at each recruiting site. Challenging cases were reviewed in multi-site team meetings and discussed with the TRACK-TBI Outcomes Core as needed to ensure consistent scoring between TRACK-TBI and TRACK-GERI.

### Statistical Methods

We summarized unweighted baseline demographic, clinical and injury characteristics overall and stratified by age group (Young-old, age 65-74 years; Middle-old, age 75-84 years; Oldest-old, age 85+ years) and by pre-injury cognitive status (unimpaired, MCI, dementia) using mean and standard deviation, frequencies and percentages. Age groups and pre-injury cognition groups were compared using Kruskal-Wallis rank sum test, Pearson’s Chi-squared test and Fisher’s exact test.

Inverse Probability Weighting (IPW)29 was used to address outcome data missingness.30,31 IPW mitigates attrition bias such that the weighted analysis is considered representative of the entire baseline cohort32 (see **Supplementary Methods** for further details). We additionally performed unweighted analyses for comparison.

We estimated 2-week and 6-month GOSE using IPW and stratified by age-group and pre-injury cognition. We summarized weighted proportions of independence at home (GOSE 5+) by age groups and pre-injury cognition and compared groups using Chi-squared tests with Rao & Scott adjustment. This analysis was repeated for survivors at 2-weeks to inform post-acute prognostication.

To further elucidate post-acute rehabilitation potential, we used the reliable change index (RCI) to estimate reliable improvement and deterioration from 2-weeks to 6-months post-injury.33,34 The RCI calculates the standardized difference between longitudinally repeated measures, adjusted for the reliability of the score and the expected change in the normative population (represented by our control group). To guide design of future trials, we calculated the minimum change required to achieve a reliable improvement or deterioration and the associated effect size (see **Supplementary Methods** for further details). We also tested the agreement between GOSE, Activities of Daily Living (ADL, Supplements) and Functional Activities Questionnaire (FAQ)’s reliable change using weighted Cohen’s kappa, and interpreted the results as proposed by Landis and Koch.35 With a 2-sided α= 0.05, power of 80% and a sample size of 154, the minimally detectable effect size for age group or cognitive status vs dichotomized GOSE is 0.25. With a 95% confidence interval (CI), the margin of error is 7.9% for a 50% proportion (CI width 15.8%) and 4.7% for a 10% or 90% proportion (CI width 9.4%). A two-sided p-value<0.05 was considered statistically significant. Statistical analyses were performed in R 4.4.2.36 The “survey” package was used for weighted analysis.

## Results

Among the 253 enrolled participants with mild TBI (mTBI cohort, **eFigure 1**), mean (SD, range) age was 77 years (8, 65-99 years, Table 1) and 129 (51%) were female. Pre-injury, 99 (51%) had unimpaired cognition (CDR=0), 68 (35%) had MCI (CDR=0.5) and 28 (15%) had dementia (CDR>0.5) (58 [23%] had data on preinjury cognition).

**Table 1.** Baseline features of older adults with TBI GCS 13-15 by age group and preinjury cognition.

| No. (%) |  |  |  |  |  |  |  |  |  |  |
| --- | --- | --- | --- | --- | --- | --- | --- | --- | --- | --- |
| Characteristic | Age group |  |  |  |  | Preinjury cognition |  |  |  |  |
|  | Overall | 65-74y | 75-84y | 85+y | p-value<sup>2</sup> | Overall | Unimpaired | MCI | Dementia | p-value<sup>2</sup> |
|  | N = 253<sup>1</sup> | N = 108<sup>1</sup> | N = 89<sup>1</sup> | N = 56<sup>1</sup> |  | N = 195<sup>1</sup> | N = 99<sup>1</sup> | N = 68<sup>1</sup> | N = 28<sup>1</sup> |  |
| **Age, mean (SD), y** | 77 (8) | 70 (3) | 79 (3) | 89 (4) | - | 78 (8) | 76 (8) | 78 (7) | 82 (9) | **0.005** |
| **Male sex** | 124 (49) | 55 (51) | 39 (44) | 30 (54) | 0.5 | 100 (51) | 48 (48) | 35 (51) | 17 (61) | 0.5 |
| **Race** |  |  |  |  | 0.3 |  |  |  |  | 0.5 |
| White | 195 (78) | 83 (78) | 66 (75) | 46 (84) |  | 153 (80) | 78 (80) | 54 (81) | 21 (75) |  |
| Black | 16 (6.4) | 10 (9.4) | 5 (6) | 1 (2) |  | 10 (5.2) | 3 (3.1) | 4 (6) | 3 (11) |  |
| Other | 38 (15) | 13 (12) | 17 (19) | 8 (14) |  | 29 (15) | 16 (16) | 9 (13) | 4 (14) |  |
| **Hispanic or Latino** | 13 (5.2) | 5 (4.7) | 4 (5) | 4 (7) | 0.8 | 9 (4.6) | 2 (2.0) | 6 (9) | 1 (4) | 0.10 |
| **Education, mean (SD), y** | 15 (3) | 15 (3) | 14 (3) | 14 (5) | 0.3 | 15 (3) | 15 (3) | 14 (3) | 14 (4) | 0.4 |
| **Employment status** |  |  |  |  | **<0.001** |  |  |  |  | **<0.001** |
| Retired | 190 (83) | 66 (68) | 74 (90) | 50 (98) |  | 152 (83) | 66 (73) | 59 (89) | 27 (100) |  |
| Employed | 40 (17) | 31 (32) | 8 (10) | 1 (2) |  | 32 (17) | 25 (27) | 7 (11) | 0 (0) |  |
| **Marital status** |  |  |  |  | **<0.001** |  |  |  |  | 0.5 |
| Married | 127 (52) | 66 (65) | 45 (52) | 16 (30) |  | 102 (53) | 55 (57) | 33 (49) | 14 (50) |  |
| Widowed | 61 (25) | 10 (9.8) | 21 (24) | 30 (56) |  | 48 (25) | 20 (21) | 17 (25) | 11 (39) |  |
| Divorced | 34 (14) | 14 (14) | 16 (19) | 4 (7) |  | 27 (14) | 13 (13) | 12 (18) | 2 (7) |  |
| Never married | 20 (8.3) | 12 (12) | 4 (5) | 4 (7) |  | 15 (7.8) | 9 (9.3) | 5 (7.5) | 1 (4) |  |
| **Living situation** |  |  |  |  | **<0.001** |  |  |  |  | **<0.001** |
| Independent, lives with others | 159 (65) | 74 (73) | 59 (67) | 26 (47) |  | 127 (65) | 64 (65) | 43 (63) | 20 (71) |  |
| Independent, lives alone | 78 (32) | 27 (26) | 29 (33) | 22 (40) |  | 62 (32) | 34 (34) | 25 (37) | 3 (11) |  |
| SNF/Nursing home | 8 (3.3) | 1 (1.0) | 0 (0) | 7 (13) |  | 6 (3.1) | 1 (1.0) | 0 (0) | 5 (18) |  |
| **Health Insurance** |  |  |  |  | 0.8 |  |  |  |  | 0.11 |
| Private insurance | 72 (30) | 31 (30) | 27 (31) | 14 (26) |  | 59 (31) | 36 (37) | 15 (22) | 8 (29) |  |
| Medicare/Medicaid | 172 (70) | 72 (70) | 60 (69) | 40 (74) |  | 134 (69) | 61 (63) | 53 (78) | 20 (71) |  |
| **Smoking status** |  |  |  |  | 0.3 |  |  |  |  | 0.3 |
| Current smoker | 16 (7.8) | 9 (11) | 6 (8.1) | 1 (2.2) |  | 13 (7.8) | 6 (7) | 5 (8.6) | 2 (8) |  |
| Past smoker | 71 (35) | 29 (34) | 29 (39) | 13 (29) |  | 54 (33) | 28 (34) | 22 (38) | 4 (16) |  |
| Never smoked | 117 (57) | 47 (55) | 39 (53) | 31 (69) |  | 99 (60) | 49 (59) | 31 (53) | 19 (76) |  |
| **Alcohol Use Disorder Identification Test (range 0-12 [worst]), mean (SD)** | 2 (2) | 2 (2) | 1 (2) | 1 (2) | 0.061 | 2 (2) | 2 (2) | 2 (2) | 2 (3) | 0.7 |
| **Family history (1<sup>st</sup> degree relative)** |  |  |  |  | 0.8 |  |  |  |  | 0.4 |
| Alzheimer's Disease/other dementia | 38 (36) | 23 (41) | 11 (31) | 4 (27) |  | 32 (38) | 20 (47) | 9 (30) | 3 (27) |  |
| Parkinson's Disease | 15 (14) | 8 (14) | 5 (14) | 2 (13) |  | 13 (15) | 6 (14) | 4 (13) | 3 (27) |  |
| None | 54 (50) | 25 (45) | 20 (56) | 9 (60) |  | 39 (46) | 17 (40) | 17 (57) | 5 (45) |  |
| **History of prior TBI** |  |  |  |  | 0.5 |  |  |  |  | 0.6 |
| More than one TBI | 4 (2.3) | 2 (2.5) | 2 (3) | 0 (0) |  | 4 (2.8) | 3 (3.8) | 0 (0) | 1 (6) |  |
| One TBI | 26 (15) | 11 (14) | 12 (20) | 3 (9) |  | 22 (15) | 13 (17) | 7 (15) | 2 (12) |  |
| None | 144 (83) | 66 (84) | 46 (77) | 32 (91) |  | 117 (82) | 62 (79) | 41 (85) | 14 (82) |  |
| **Number of comorbidities, mean (SD)** | 5 (3) | 4 (3) | 5 (3) | 5 (2) | **0.024** | 5 (3) | 4 (3) | 5 (3) | 5 (2) | 0.2 |
| **Number of medications, mean (SD)** | 15 (10) | 14 (12) | 15 (10) | 14 (8) | 0.6 | 14 (10) | 13 (10) | 16 (13) | 14 (7) | 0.3 |
| **Anticoagulants** | 55 (22) | 17 (16) | 21 (24) | 17 (33) | **0.047** | 43 (22) | 21 (22) | 13 (19) | 9 (32) | 0.4 |
| **Platelet aggregation inhibitors** | 72 (30) | 28 (27) | 30 (36) | 14 (27) | 0.4 | 62 (34) | 28 (30) | 26 (40) | 8 (30) | 0.4 |
| **Pre-injury cognition** |  |  |  |  | **<0.001** |  |  |  |  | - |
| Unimpaired | 99 (51) | 48 (62) | 31 (43) | 20 (43) |  | 99 (51) | 99 (100) | - | - |  |
| MCI | 68 (35) | 22 (29) | 34 (47) | 12 (26) |  | 68 (35) | - | 68 (100) | - |  |
| Dementia | 28 (14) | 7 (9) | 7 (10) | 14 (30) |  | 28 (14) | - | - | 28 (100) |  |
| **ADL (Activities of Daily Living, range 1-27 [worst]), mean (SD)** | 4 (5) | 2 (5) | 3 (4) | 7 (7) | **<0.001** | 4 (6) | 2 (4) | 5 (5) | 10 (8) | **<0.001** |
| **GFI (Groningen Frailty Indicator, range 0-15 [worst]), mean (SD)** | 4 (3) | 4 (3) | 4 (2) | 6 (3) | **0.016** | 4 (3) | 4 (2) | 4 (2) | 7 (3) | **<0.001** |
| **Injury mechanism** |  |  |  |  | **0.002** |  |  |  |  | 0.2 |
| Ground level fall | 152 (61) | 54 (50) | 55 (63) | 43 (77) |  | 114 (58) | 56 (57) | 42 (62) | 16 (57) |  |
| Fall from height | 45 (18) | 19 (18) | 16 (18) | 10 (18) |  | 41 (21) | 17 (17) | 15 (22) | 9 (32) |  |
| Motor vehicle accident | 45 (18) | 27 (25) | 15 (17) | 3 (5.4) |  | 32 (16) | 23 (23) | 7 (10) | 2 (7.1) |  |
| Other | 9 (3.6) | 8 (7.4) | 1 (1.1) | 0 (0) |  | 8 (4.1) | 3 (3.0) | 4 (5.9) | 1 (3.6) |  |
| **Glasgow Coma Scale** |  |  |  |  | >0.9 |  |  |  |  | 0.13 |
| 15 | 179 (71) | 74 (69) | 64 (72) | 41 (73) |  | 134 (69) | 68 (69) | 52 (76) | 14 (50) |  |
| 14 | 62 (25) | 28 (26) | 22 (25) | 12 (21) |  | 50 (26) | 26 (26) | 13 (19) | 11 (39) |  |
| 13 | 12 (4.7) | 6 (5.6) | 3 (3.4) | 3 (5.4) |  | 11 (5.6) | 5 (5.1) | 3 (4.4) | 3 (11) |  |
| **LOC** | 104 (42) | 52 (49) | 37 (43) | 15 (28) | **0.040** | 82 (43) | 51 (53) | 23 (35) | 8 (30) | **0.025** |
| **PTA** | 114 (46) | 55 (51) | 39 (45) | 20 (37) | 0.2 | 91 (48) | 50 (52) | 29 (44) | 12 (44) | 0.6 |
| **AOC** | 97 (40) | 47 (44) | 29 (34) | 21 (39) | 0.4 | 79 (42) | 41 (43) | 28 (42) | 10 (37) | 0.9 |
| **TBI symptoms (LOC/PTA/AOC)** | 154 (63) | 73 (69) | 53 (62) | 28 (52) | 0.11 | 121 (64) | 63 (66) | 42 (64) | 16 (59) | 0.8 |
| **Extracranial injury** | 116 (46) | 50 (47) | 43 (49) | 23 (42) | 0.7 | 96 (50) | 45 (47) | 35 (51) | 16 (57) | 0.6 |
| **Injury Severity Score, mean (SD)** | 12 (10) | 12 (10) | 13 (11) | 12 (9) | 0.8 | 13 (11) | 14 (11) | 11 (7) | 17 (17) | 0.2 |
| **CT-positive mTBI** | 145 (57) | 61 (56) | 51 (57) | 33 (59) | >0.9 | 117 (60) | 61 (62) | 39 (57) | 17 (61) | 0.9 |
| **Pupil reactivity** | 247 (100) | 104 (100) | 89 (100) | 54 (100) |  | 190 (100) | 98 (100) | 65 (100) | 27 (100) |  |
| **ED hemoglobin (mg/dL), mean (SD)** | 13.16 (1.78) | 13.54 (1.55) | 13.00 (1.83) | 12.75 (1.98) | **0.013** | 13.11 (1.82) | 13.14 (1.58) | 13.14 (2.03) | 12.95 (2.14) | 0.8 |
| **ED blood glucose (mg/dL), mean (SD)** | 131 (56) | 136 (67) | 130 (47) | 124 (46) | 0.6 | 129 (50) | 129 (49) | 126 (33) | 133 (81) | 0.3 |
| **Hypotension (SBP < 90 mmHg)** | 18 (7.1) | 8 (7.4) | 7 (7.9) | 3 (5.4) | >0.9 | 10 (5.1) | 4 (4.0) | 5 (7.4) | 1 (3.6) | 0.7 |
| **Hypoxia (SpO2 < 90)** | 16 (6.3) | 6 (5.6) | 7 (7.9) | 3 (5.4) | 0.8 | 14 (7.2) | 5 (5.1) | 5 (7.4) | 4 (14) | 0.3 |
| **ED disposition** |  |  |  |  | 0.5 |  |  |  |  | >0.9 |
| ED Discharge | 52 (21) | 24 (22) | 19 (21) | 9 (16) |  | 40 (21) | 22 (22) | 14 (21) | 4 (14) |  |
| Hospital admit no ICU | 126 (50) | 52 (48) | 48 (54) | 26 (46) |  | 92 (47) | 46 (46) | 32 (47) | 14 (50) |  |
| Hospital admit with ICU | 75 (30) | 32 (30) | 22 (25) | 21 (38) |  | 63 (32) | 31 (31) | 22 (32) | 10 (36) |  |
| **Discharge disposition** |  |  |  |  | 0.3 |  |  |  |  | 0.5 |
| Home | 173 (71) | 82 (78) | 57 (66) | 34 (64) |  | 132 (70) | 66 (70) | 48 (73) | 18 (64) |  |
| Other hospital, SNF or rehabilitation unit | 58 (24) | 18 (17) | 25 (29) | 15 (28) |  | 45 (24) | 23 (24) | 16 (24) | 6 (21) |  |
| LTAC/Nursing home | 4 (1.6) | 1 (1) | 2 (2) | 1 (1.9) |  | 3 (1.6) | 2 (2.1) | 0 (0) | 1 (4) |  |
| Died in hospital | 10 (4.1) | 4 (4) | 3 (3) | 3 (5.7) |  | 8 (4.3) | 3 (3.2) | 2 (3.0) | 3 (11) |  |

<sup>1</sup>Mean (SD); n ()
<sup>2</sup>Comparison of age/cognition groups: Kruskal-Wallis rank sum test; Pearson's Chi-squared test; Fisher's exact test
**Abbreviations:** AIS, Abbreviated Injury Scale; AOC, Alteration of Consciousness; CT, computerized tomography; ED, Emergency Department; GCS, Glasgow Coma Scale; ISS, Injury Severity Scale; LOC, Loss of Consciousness; LTAC, Long-Term Acute Care facility; PTA, Post-Traumatic Amnesia; SBP, Systolic Blood Pressure; SNF, Specialized Nursing Facility; SPO2, blood Oxygen Saturation; TBI, Traumatic Brain Injury; y, years.
**Missing data (n, %):** Race (4, 1.6%), Hispanic or Latino (2, 0.8%), Years of education (13, 5.1%), Employment status (23, 9.1%), marital status (11, 4.3%), living situation (8, 3.2%), health insurance (9, 3.6%), smoking status (49, 19.4%), Alcohol use disorder identification test (24, 9.5%), family history (146, 58%), history of prior TBI (79, 31%), number of comorbidities (3, 1.2%), anticoagulants (7, 2.8%), platelet aggregation inhibitors (15, 5.9%), pre-injury cognition (58, 23%), ADL (22, 8.7%), GFI (45, 18%), injury mechanism (2, 0.8%), LOC (6, 2.4%), PTA (6, 2.4%), AOC (8, 3.2%), TBI symptoms (8, 3.2%), extracranial injury (3, 1.2%), ISS (51, 20%), pupil reactivity (6, 2.4%), ED hemoglobin (12, 4.7%), ED blood glucose (13, 5.1%), discharge disposition (8, 3.2%).

Demographic, pre-injury, and injury characteristics of our cohort (N=253), stratified by age-group and pre-injury cognition are reported in **Table 1**. Anti-coagulant use increased with age but not platelet aggregation inhibitor use. Injury mechanism of ground level fall was increasingly prevalent with increasing age (44%, 57%, 73%; *P*=0.004), but was not associated with pre-injury cognitive status (57%, 62%, 57%, *P*=0.2). The oldest-old and those with pre-injury MCI/dementia were the least likely to report loss of consciousness (49%, 43%, 28%, *P*=0.04; 53%, 35%, 30%, *P*=0.025). Other injury characteristics were similar across groups.

At 6 months, 154 (61%) had GOSE assessed (follow-up group). Those who were lost to follow up (drop-out group, n=99, 39%) had lower mean (SD) years of education, (14 [4] years vs 15 [3] years in the follow-up group, *P*=0.03), lower mean (SD) injury severity score (10 [7] vs 14 [12], *P*=0.03), and their study partner’s race was less likely to be Caucasian (56% vs 75%, *P*=0.004). All other demographic, clinical, functional, injury and study partner characteristics by drop-out vs follow-up were similar (**eTable 1**).

73% of older adults achieved at least independence at home (GOSE 5+) by 6-months post-injury. GOSE varied significantly by age and pre-injury cognition with higher proportion of home independence observed in younger age-category (84% in younger-old, 74% in middle-old and 52% in oldest-old, *P*=0.004) or better pre-injury cognitive status (86% in cognitively-unimpaired, 73% in MCI and 23% in dementia; *P*<.001; **Figure 1**).

**Figure 1.**
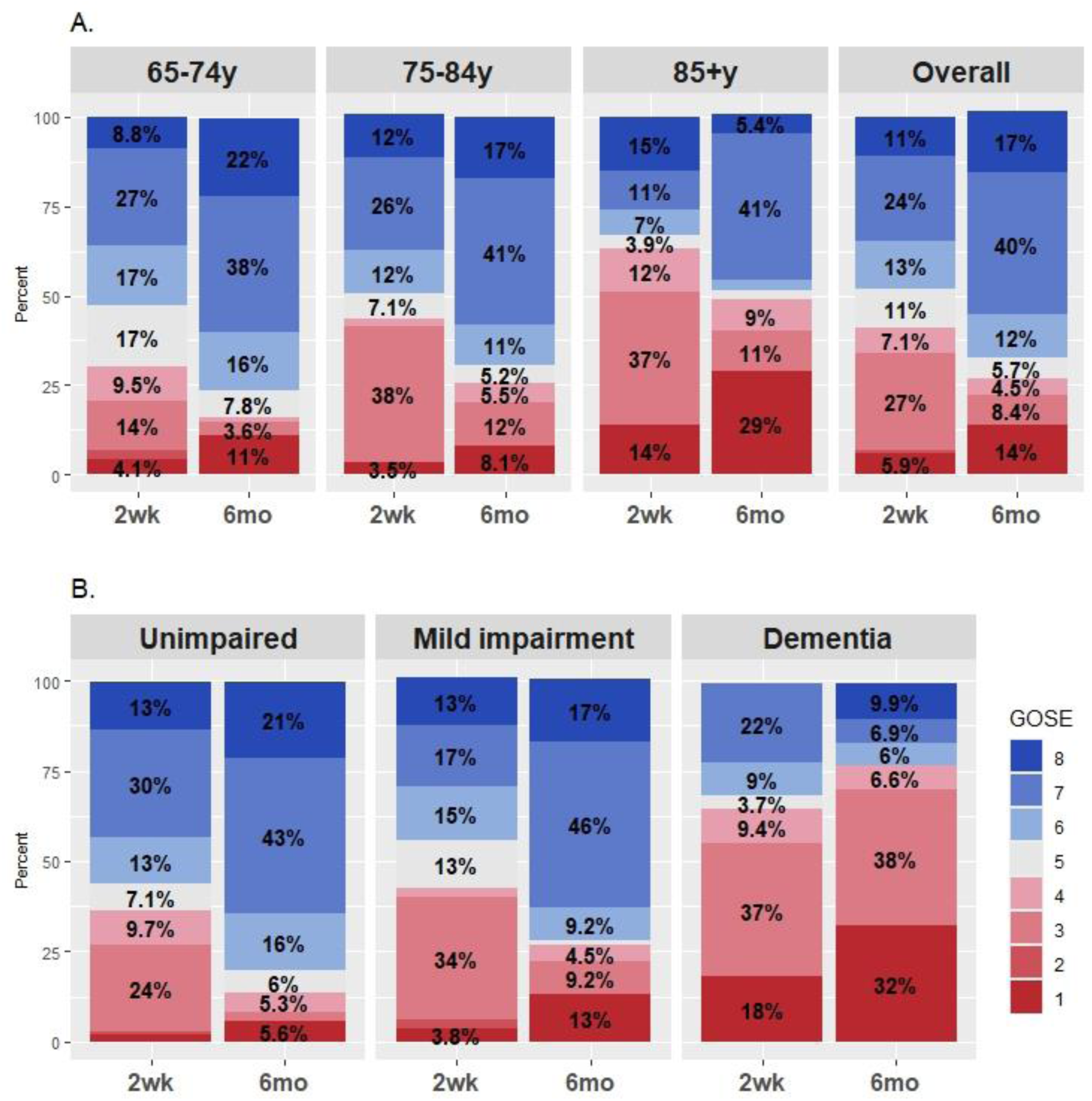
Weighted 6-month GOSE by age group and pre-injury cognition. Distribution of GOSE at 2 weeks (2wk) and 6 months (6mo) is shown stratified by age group in panel A and by preinjury cognition in panel B. Percentages lower than 3% are not labelled for figure clarity. GOSE, Glasgow Outcome Scale Extended; Mild impairment, Mild cognitive impairment; Unimpaired, cognitively unimpaired; y, years old.

All age and preinjury cognition groups showed an increase in proportion of independence at home from 2-weeks to 6-months, except for those with dementia, who got worse (35% independent at 2-weeks vs 23% at 6 months). GOSE trajectories are illustrated in **Figure 2A**.

**Figure 2.**
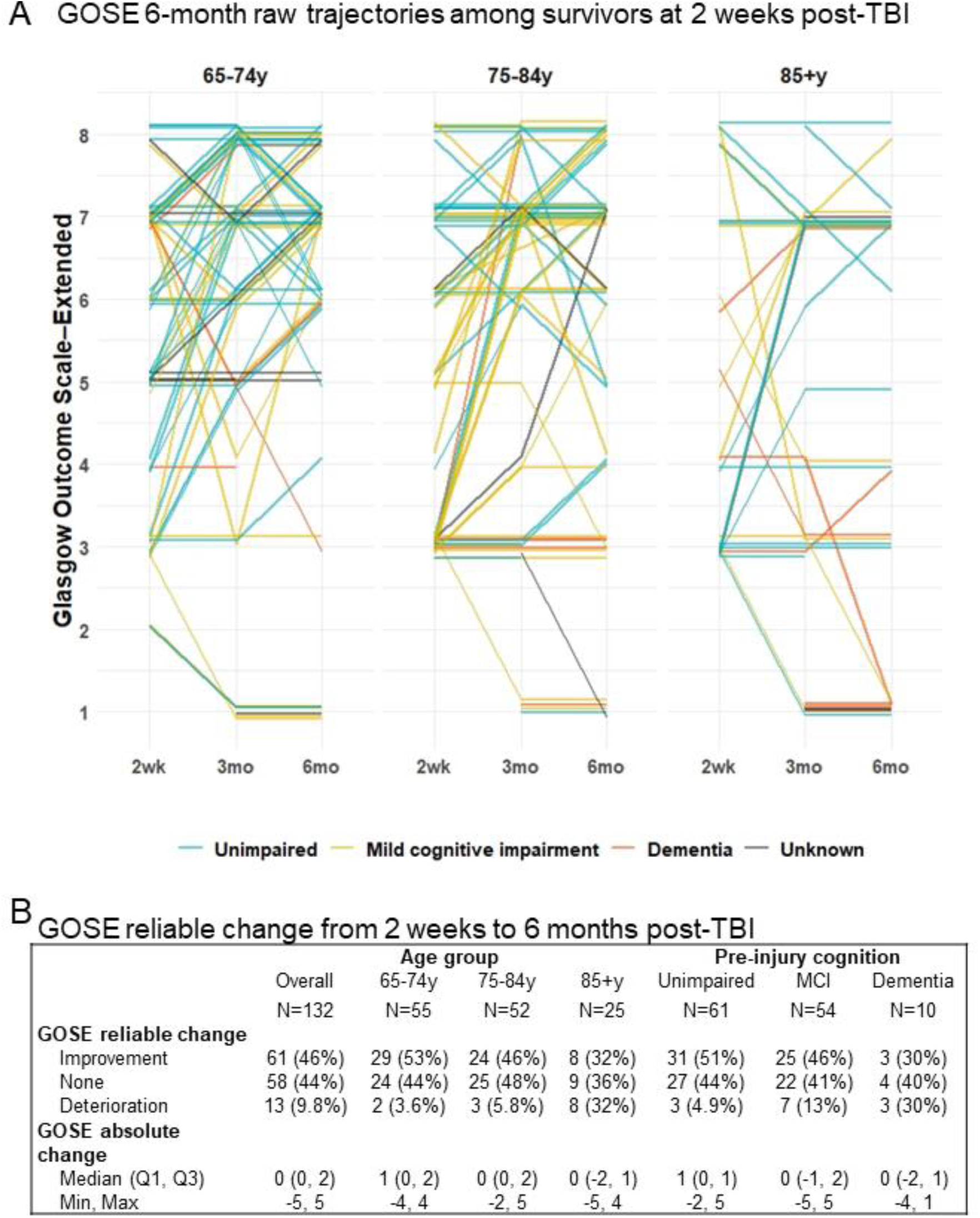
Glasgow Outcome Scale-Extended 6-month trajectories and reliable change. A. GOSE trajectories of survivors at 2 weeks who had GOSE assessed more than once are shown stratified by age-category and color-coded by pre-injury cognitive status: 65-74 years old (n=69), 75-84 years old (n=65) and 85+ years old (n=35). Cognitively unimpaired are shown in teal; mild cognitive impairment, yellow; dementia, orange. Each line represents an individual participant. B. GOSE reliable and absolute change from 2 weeks to 6 months, by age category and pre-injury cognitive status, for participants who had both 2-week and 6-month GOSE available. Abbreviations: 2wk, 2 weeks; 3mo, 3 months; 6mo, 6 months; y, years old.

Overall, full recovery rate was 11% at 2-weeks and 17% at 6-months; mortality rate was 5.9% and 14%, respectively. 2-week mortality was 3.4-fold among the oldest-old vs younger-old and 9.5-fold among those with pre-injury dementia vs unimpaired cognition (2.6-fold and 5.7-fold at 6-months). Among survivors at 2-weeks, similarly, full recovery was achieved by 18% overall, 25% of the younger-old and 6.7% of the oldest-old. 6-month mortality rate was 4.4%, with 9.3-fold rate among the oldest-old compared to the young-old and 6-fold in those with pre-injury dementia vs cognitively-unimpaired. (**Table 2).**

**Table 2.** Weighted 2-week and 6-month Glasgow Outcome Scale Extended by age group and pre-injury cognition.

|  | Age group |  |  |  |  | Pre-injury cognition |  |  |  |
| --- | --- | --- | --- | --- | --- | --- | --- | --- | --- |
|  | Overall | 65-74y | 75-84y | 85+y | p-value<sup>1</sup> | Unimpaired | MCI | Dementia | p-value<sup>1</sup> |
| **2-week GOSE, %** |  |  |  |  | **0.004** |  |  |  | 0.094 |
| 1 | 5.9 | 4.1 | 3.5 | 14 |  | 1.9 | 3.8 | 18 |  |
| 2 | 1.1 | 2.6 | 0 | 0 |  | 0.9 | 2.4 | 0 |  |
| 3 | 27 | 14 | 38 | 37 |  | 24 | 34 | 37 |  |
| 4 | 7.1 | 9.5 | 2.1 | 12 |  | 9.7 | 2.5 | 9.4 |  |
| 5 | 11 | 17 | 7.1 | 3.9 |  | 7.1 | 13 | 3.7 |  |
| 6 | 13 | 17 | 12 | 7.0 |  | 13 | 15 | 9.0 |  |
| 7 | 24 | 27 | 26 | 11 |  | 30 | 17 | 22 |  |
| 8 | 11 | 8.8 | 12 | 15 |  | 13 | 13 | 0 |  |
| **2-week GOSE 5+, %** | 59 | 70 | 57 | 37 | **0.008** | 63 | 58 | 35 | 0.10 |
| **6-month GOSE, %** |  |  |  |  |  |  |  |  |  |
| 1 | 14 | 11 | 8.1 | 29 |  | 5.6 | 13 | 32 |  |
| 2 | 0 | 0 | 0 | 0 |  | 0 | 0 | 0 |  |
| 3 | 8.4 | 3.6 | 12 | 11 |  | 2.7 | 9.2 | 38 |  |
| 4 | 4.5 | 1.2 | 5.5 | 9.0 |  | 5.3 | 4.5 | 6.6 |  |
| 5 | 5.7 | 7.8 | 5.2 | 2.5 |  | 6.0 | 1.4 | 0 |  |
| 6 | 12 | 16 | 11 | 2.9 |  | 16 | 9.2 | 6.0 |  |
| 7 | 40 | 38 | 41 | 41 |  | 43 | 46 | 6.9 |  |
| 8 | 17 | 22 | 17 | 5.4 |  | 21 | 17 | 9.9 |  |
| **6-month GOSE 5+, %** | 73 | 84 | 74 | 52 | **0.004** | 86 | 73 | 23 | **<0.001** |
| **6-month GOSE for survivors at 2 weeks, %** |  |  |  |  | 0.067 |  |  |  | **<0.001** |
| 1 | 4.4 | 1.4 | 3.4 | 13 |  | 1.3 | 6.5 | 7.8 |  |
| 3 | 9.3 | 4.0 | 13 | 13 |  | 2.9 | 9.9 | 52 |  |
| 4 | 5.0 | 1.4 | 5.8 | 11 |  | 5.5 | 4.8 | 9.0 |  |
| 5 | 6.4 | 8.7 | 5.5 | 3.1 |  | 6.3 | 1.5 | 0 |  |
| 6 | 13 | 18 | 12 | 3.5 |  | 17 | 9.9 | 8.1 |  |
| 7 | 44 | 42 | 43 | 50 |  | 45 | 49 | 9.4 |  |
| 8 | 18 | 25 | 17 | 6.7 |  | 22 | 18 | 13 |  |
| **6-month GOSE 5+ for survivors at 2 weeks, %** | 81 | 93 | 78 | 63 | **0.004** | 90 | 79 | 31 | **<0.001** |

<sup>1</sup> Pearson's X<sup>2</sup>: Rao & Scott adjustment
Abbreviations: GOSE, Glasgow Coma Scale-Extended; MCI = mild cognitive impairment; y = years.
GOSE score: 1- dead, 2- vegetative state, 3- lower severe disability, 4- upper severe disability, 5- lower moderate disability, 6-upper moderate disability, 7- lower good recovery, 8-upper good recovery. GOSE 5+ indicates functional independence at home.

Reliable change in GOSE between 2 weeks and 6 months in survivors at 2 weeks showed that overall, deterioration occurred in 9.8% of participants (3.6% in the younger-old, 5.8% in middle-old and 32% in the oldest-old, **Figure 2B**). Reliable improvement was seen in 46% overall, 53% of the younger-old, 46% in middle-old and 32% in the oldest-old. Pre-injury cognitive status groups showed similar percentages, parallel to age category groups (**Figure 2B**). Reliable changes in ADL, FAQ and CDR are reported in **eTable 2**.

The minimal score change and effect sizes required to detect a reliable improvement or deterioration with GOSE, ADL and FAQ are presented in **Table 3**. Compared to ADL and FAQ, GOSE was the measure that captured the largest proportion of reliable improvement (46% with GOSE vs 33% with ADL and 18% with FAQ) and reliable deterioration (10% vs 5.8% and 5.2%). Cohen’s weighted kappa for agreement between GOSE and ADL was fair (n=108; weighted κ=0.39, 95% CI: 0.24-0.55); and same for GOSE and FAQ (n=103; weighted κ=0.25, 95% CI 0.-0.41).

**Table 3.** Minimal reliable score change and effect sizes for functional outcomes.

| **Assessment** | **Reliable improvement, points (raw)** | **Reliable deterioration, points (raw)** | **Effect size for reliable improvement** | **Effect size for reliable deterioration** |
| --- | --- | --- | --- | --- |
| GOSE | +1 (+0.68) | -2 (-1.8) | 0.52 | 1.1 |
| ADL | +4 (+3.9) | -4 (-3.1) | 0.84 | 0.67 |
| FAQ | +5 (+4.8) | -5 (-4.5) | 1 | 1.01 |
| Abbreviations: ADL, Activities of daily living; FAQ, Functional activities questionnaire; GOSE, Glasgow outcome scale- extended. |  |  |  |  |

## Discussion

In this prospective study we report 6mo GOSE, functional, geriatric, cognitive and TBI CDE outcomes in a population of real-world older adults after mild TBI, by age group and pre-injury cognition. Younger and cognitively-unimpaired participants had better outcomes. Additionally, all age and cognition groups showed higher independence at home at 6-months as compared to 2-weeks with the exception of those with pre-injury dementia who showed progressive decline rather than recovery. Loss of consciousness was less often documented in the oldest-old and those with cognitive impairment despite similar injury severity by other metrics, suggesting under-recognition, under-reporting, or different TBI pathophysiology in these groups.

Loss to follow-up was moderate (39%), and was addressed by IPW. This loss to follow-up rate is substantially lower than that among older adults who participated in CENTER-TBI, a large multi-center European cohort study (56%)37 and comparable to that reported among all-ages adults (≥17 years) with mTBI who participated in TRACK-TBI, a large multi-center U.S. cohort study (37%).38 A systematic review found an average drop-out rate of 9% at 6-months in observational all-severity all-age TBI studies, although it varied markedly (range: 0%-75%).30 In our study, adherent participants had more years of education, were more likely to have MCI (vs no cognitive impairment or dementia) and there were indications for possible better study partner availability. This underscores the inherent challenges of longitudinal TBI research in older populations and the need for research methods optimization to support age-inclusive TBI research, specifically reducing study partner burden, which is a modifiable factor.

Most prior studies of mTBI in older adults excluded those with pre-existing cognitive impairment or a high-burden of pre-existing comorbidities and, unsurprisingly, reported more optimistic outcomes than our cohort. A meta-analysis of mTBI in older adults found that 67% achieved good recovery (GOSE=7-8)39 compared to 57% in our cohort (60% and 64% in the young-old and those with no cognitive- impairment, respectively); Other studies37,40 found 44%-53% complete recovery (GOSE=8), compared to 17% in our cohort (22%; 21%).

6-month mTBI mortality rate in older adults was previously reported as 9.4%41 vs our finding of 14% (11%; 5.6%). A meta-analysis of in-hospital mortality after mTBI in older adults reports 5% mortality,42 similarly to our 2-week mTBI mortality rate of 5.9% (4.1%; 1.9%). In our study, weighted 6-month mortality among young-, middle- and oldest-old with mTBI was 11%, 8.1% and 29% compared to expected 6-month mortality in the general older adult population of 1.8%, 4.3% and 14.3%, respectively.43 Together, these data highlight the expected age-related increase in mortality that is totally independent of TBI while also allowing for a rough estimate of the specific excess TBI-related mortality across age-categories in our study: 9%, 3.8% and 14.7%, respectively. Of course, prevalence of pre-existing dementia increased with increasing age in our cohort. Thus, consideration of the expected versus excess mortality risk associated with dementia warrants consideration as well. Expected mortality risk in persons with dementia is estimated to be 2-fold compared to persons with no cognitive impairment,44–46 but was 5.7-fold in our cohort of older adults with mTBI. Therefore, considering expected baseline mortality risks and the excess risk conferred by TBI, dementia status appears to be a far more substantial modifier of post-TBI mortality risk, compared with age.

Interestingly, and encouragingly, the proportion of persons with MCI with GOSE 5+ increased from 58% at 2-weeks to 73% at 6-months, which is comparable to overall recovery across the entire cohort and unlike those with dementia who deteriorated. While this may partly reflect the fact that pre-injury lower-functioning individuals have less ground to cover to close the gap to their pre-injury level, it also suggests that individuals with pre-injury MCI retain very similar 6-month recovery potential to individuals without preinjury cognitive impairment.

Comparison of GOSE, ADL and FAQ revealed limited agreement, highlighting their complementary constructs. GOSE was the most sensitive to reliable change, but may have limitations in older adults. GOSE scores are relational to pre-injury function, so between-patient comparisons are not straightforward, particularly given the heterogeneity of preinjury function in older adults. In persons with pre-existing disability who regained most of their baseline function, residual limitation may not be captured by GOSE. In addition, GOSE items regarding return to work are not applicable for many older adults. ADL’s basic functions such as bathing and walking, or FAQ’s cognitively-mediated functions such as managing finances or remembering appointments and medications may be more relevant, and complementary to GOSE.

Other studies have estimated GOSE’s correlation to TBI outcome measures in all-age all-severity TBI cohorts: neurocognitive measures (mean ρ=0.21) and self-report outcomes (mean ρ=0.49),47 and life satisfaction (ρ=0.38),48 finding medium-low associations. GOSE=8 corresponded well to other outcomes measures, but some patients classified as GOSE=7 showed impairment in multiple other measures.47 The Functional Status Examination was shown to yield more information than the GOSE, across a wide range of functional status.49 These findings highlight the need for multiple measures to capture TBI’s multi-dimensional impact.

Our study has many strengths. It presents real-world evidence on an inclusive cohort of older adults. We provide rich data on patients’ characteristics and geriatric functional outcomes, and hope it will support future studies in this population. However, our cohort does not represent those who presented to non-trauma centers or not at all. The study also excluded participants with moderate-severe TBI, due to the insufficient sample sizes (n=9 and n=6, respectively). Further research on moderate-severe TBI in older adults is warranted. The use of a dichotomized GOSE (GOSE 5+) is prevalent in the literature due to its simplicity and clinical relevance, but results in loss of information and may introduce bias.28 For this reason, we additionally report the full GOSE.

## Conclusions

Real-world 6-month functional outcomes of older adults after mild TBI show better outcomes for younger-old and those without pre-injury dementia. GOSE effectively detects reliable functional change after mild TBI in older adults, and ADL and FAQ capture additional non-overlapping domains of function important for this population. There is now an urgent need to apply the available observational evidence to design and execute interventional studies to improve outcome in older adults with TBI who comprise a large and rapidly expanding proportion of the global burden of this disease.

## Data sharing

Data is available on request from the corresponding author.

## Data Availability

All data produced in the present study are available upon reasonable request to the corresponding author.

