## Supplemental material for "6-Month Recovery after Mild Traumatic Brain Injury in Older Adults: A TRACK-GERI Study"

#### eMethods

##### Data collected

Demographics included age, sex, race, ethnicity, smoking status, employment status, marital status, living situation, and health insurance. Medical history included the number of comorbidities, the number of prescription medications taken pre-injury, pre-injury use of anticoagulants and platelet aggregation inhibitors, family history of dementia or Parkinson's disease in first-degree relatives, and prior TBI. We calculated a modified (e.g., missing diabetes complications and moderate-severe liver disease) Charlson Comorbidity Index (CCI).[1] Alcohol use was assessed using the Alcohol Use Disorder Identification Test (AUDIT-C).[2] We collected data on injury mechanism, TBI symptoms of loss of consciousness (LOC), post-traumatic amnesia (PTA), and alteration of consciousness (AOC), acute intracranial trauma on initial head CT (defined according to expert consensus TBI neuroimaging common data elements),[3] extracranial injury (defined as non-head Abbreviated Injury Scale [AIS] ≥2, which indicates a non-head injury that would justify a hospitalization in its own right), Injury Severity Scale (ISS), and ED disposition (discharged from the ED, admitted to ward, admitted to ICU). Clinical characteristics on arrival to the ED included GCS, pupil reactivity (bilateral, unilateral, none), hypotension (systolic blood pressure < 90 mm Hg), hypoxia (blood oxygen saturation < 90%), blood glucose and hemoglobin.

##### Inverse Probability weighting

Inverse Probability Weighting (IPW)[4] was used to address primary outcome data missingness. IPW mitigates attrition bias such that the weighted analysis is considered representative of the entire baseline cohort.[5,6] We compared those with available 6-month GOSE (follow-up group, N=154) to those missing 6-month GOSE (drop-out group, N=99) across demographic, clinical, injury, and study partner characteristics to identify missingness correlates for inclusion in the IPW model. Variables with a standardized difference greater than 0.1 between follow-up and drop-out groups were used for the model,[7] excluding those with more than 20 missingness. Next, a boosted logistic regression model was used to predict the probability of missingness.[8] The model included: study site, age, race, years of education, preinjury cognitive status (unimpaired, MCI, dementia, or unknown because study partner unavailable to complete CDR interview), ED disposition (discharge, ward admission, ICU admission). The inverse of the model's predicted probabilities were used as weights in 6-month GOSE analyses. We additionally performed unweighted analyses for comparison.

##### Reliable Change Index

To determine which functional outcome(s) most effectively capture a clinically meaningful and reliable change in functional status from 2-weeks to 6-months post-injury, we used the reliable change index (RCI).[9,10] The RCI calculates the standardized difference between longitudinally repeated measures, adjusted for the reliability of the score and the expected change in our control group population (full description of methods and results in supplements). We computed RCI for each participant and each outcome using the formula:

$$\mathrm{RCI} = \frac{\left((\mathrm{X}_2 - \mathrm{X}_1) - (\mathrm{M}_2 - \mathrm{M}_1)\right)}{SED}$$

where $\mathrm{X}_1$=participant's score at 2 weeks, $\mathrm{X}_2$=participant's score at 6 months, $\mathrm{M}_1$=mean score for the cognitively-unimpaired control group at baseline, $\mathrm{M}_2$=mean score for cognitively-unimpaired controls at 6 months, to correct for changes in scores that may occur during 6 months in the normative corresponding population. $SED$ = Standard error of the difference was calculated with the formula:

$$SED = SD_1\sqrt{2(1-r)}$$

where $SD_1$=standard deviation of 2-week score, $r$ =score's reliability coefficient, as reported in the literature. Reliability coefficients were derived from published validation studies for GOSE ($r$ =0.92)[15] and FAQ ($r$ =0.95).[12] Our ADL instrument was comprised of ADL and mobility questions, similar to the Barthel Index, and thus we relied on the published reliability of the Barthel Index ($r$ =0.97).[11] A reliable change was defined by setting α at 0.10 (two-tailed) and a 90 confidence interval. For an assessment where higher scores are better (e.g. GOSE), RCI values exceeding +1.645 were defined as "reliable improvement" and RCI values below −1.645 were defined as "reliable deterioration".[9] The opposite was true for assessments where lower scores are better (e.g., ADL). We described the reliable changes and absolute changes in GOSE, ADL, and FAQ between 2 weeks and 6 months stratified by age group and pre-injury cognitive status, with median (), frequencies, and percentages (**eTable2**). To adjust the RCI for normative changes in scores during 6 months, the difference between mean score at baseline and at 6 months was calculated for unimpaired cognition controls (n=52). The change in means for GOSE, ADL and FAQ in the control group was -0.55, 0.44 and 0.18 points, respectively.

### Activities of Daily Living (ADL) assessment form

**ADLs and Mobility**

The purpose of this next set of questions is to find out about [your/patient name's] everyday activities at home. I'm going to ask you whether [you/patient name] need(s) help from another person to complete these tasks. I'm asking about family members, friends, and home care workers.

At the present time, do/does [you/patient name] have difficulty with, need help from another person, or were unable to do any of the following:

| | DK | Normal | Has difficulty, but does by self | Requires assistance | Dependent/ Unable to do |
|---|---|---|---|---|---|
| 1. Bathing (washing and drying whole body) | 8 | 0 | 1 | 2 | 3 |
| 2. Dressing (putting on a shirt or shoes, buttoning, zippering) | 8 | 0 | 1 | 2 | 3 |
| 3. Eating (holding a fork, cutting food, drinking from a glass) | 8 | 0 | 1 | 2 | 3 |
| 4. Using the toilet (including getting on and off the toilet) | 8 | 0 | 1 | 2 | 3 |
| 5. Getting in and out of a chair | 8 | 0 | 1 | 2 | 3 |
| 6. Walking across the room | 8 | 0 | 1 | 2 | 3 |
| 7. Walking several hundred yards (e.g., a few city blocks) | 8 | 0 | 1 | 2 | 3 |
| 8. Walking up 10 steps | 8 | 0 | 1 | 2 | 3 |
| 9. Doing laundry or housework (washing the dishes, taking out the trash, scrubbing the floors) | 8 | 0 | 1 | 2 | 3 |

At the present time...how far do/does [you/patient name] walk on an average day? (12 city blocks=1 mile)
Blocks:_________ (None or less than one block = 0, DK = 888)

At the present time...do/does [you/patient name] use any of the following devices to help them walk and/or get around?
a. cane b. walker c. wheelchair d. other:______________ e. DK f. N/A

**Self-rated Health Questions**

1. Currently…would you say [your/patient's name] health was excellent, very good, good, fair, or poor?
   a. Excellent b. Very good c. Good d. Fair e. Poor f. DK

End Time:______________

#### Supplemental Figures

##### eFigure 1. CONSORT diagram

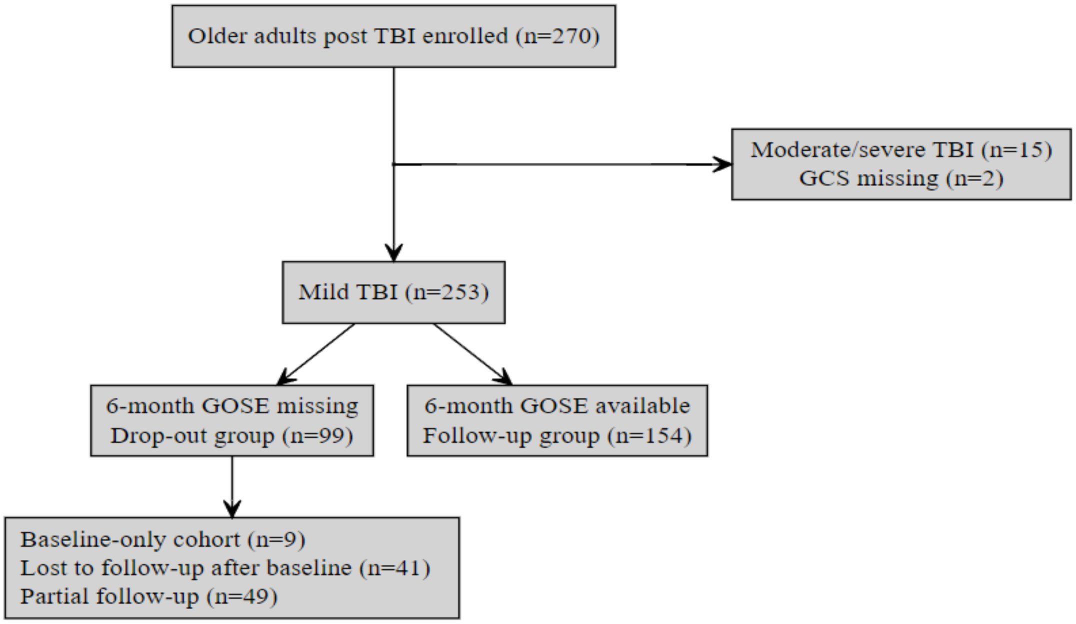

**eFigure 1.** Flow diagram of participants' disposition and inclusion in present analysis. Of 270 enrolled participants, 253 met inclusion criteria of available emergency department arrival Glasgow Coma Scale (GCS) data and sustaining mild traumatic brain injury (TBI). Of them, 154 had Glasgow Outcome Scale Extended (GOSE) assessed at 6 months. The entire cohort (n=253) was used use estimate 6-month GOSE outcomes, using inverse probability weighting.

#### Supplemental Tables

##### eTable 1. Baseline characteristics of TBI participants with versus without 6-month GOSE available for analysis

| | No. (%) | | | |
|---|---|---|---|---|
| | Missing data (n, %) | Drop-out N = 99[1] | Follow-up N = 154[1] | p-value[2] |
| **Study site** | 0 (0) | | | 0.076 |
| Site 1 | | 70 (71) | 92 (60) | |
| Site 2 | | 29 (29) | 62 (40) | |
| **Age, mean (SD), y** | 0 (0) | 78 (9) | 77 (8) | >0.9 |
| **Male gender** | 0 (0) | 43 (43) | 81 (53) | 0.2 |
| **Race** | 4 (1.6) | | | 0.093 |
| Black | | 10 (10) | 6 (4.0) | |
| Other | | 17 (17) | 21 (14) | |
| White | | 71 (72) | 124 (82) | |
| **Hispanic or Latino** | 1 (0.4) | 5 (5.1) | 8 (5.2) | >0.9 |
| **Education, mean (SD), y** | 13 (5.1) | 14 (4) | 15 (3) | **0.032** |
| **Employment status** | 23 (9.1) | | | 0.5 |
| Retired | | 73 (85) | 117 (81) | |
| Employed | | 13 (15) | 27 (19) | |
| **Marital status** | 11 (4.3) | | | 0.6 |
| Married | | 44 (48) | 83 (55) | |
| Widowed | | 24 (26) | 37 (25) | |
| Divorced | | 13 (14) | 21 (14) | |
| Never married | | 10 (11) | 10 (6.6) | |
| **Living situation** | 8 (3.2) | | | >0.9 |
| Independent- lives with others | | 61 (66) | 98 (64) | |
| Independent- lives alone | | 28 (30) | 50 (33) | |
| SNF/Nursing home | | 3 (3.3) | 5 (3.3) | |
| **Health Insurance** | 9 (3.6) | | | 0.2 |
| Private insurance | | 23 (25) | 49 (32) | |
| Medicare/Medicaid | | 69 (75) | 103 (68) | |
| **Smoking status** | 49 (19) | | | 0.5 |
| Current smoker | | 8 (10) | 8 (6.3) | |
| Past smoker | | 24 (31) | 47 (37) | |
| Never smoked | | 45 (58) | 72 (57) | |
| **Alcohol Use Disorder Identification Test (range 0-12 (worst)), mean (SD)** | 24 (9.5) | 2 (2) | 1 (2) | 0.7 |
| **Family history (1st degree relatives)** | 146 (58) | | | 0.4 |
| Alzheimer's Disease/other dementia | | 11 (32) | 27 (37) | |
| Parkinson's Disease | | 7 (21) | 8 (11) | |

| | | | | |
|---|---|---|---|---|
| None | | 16 (47) | 38 (52) | |
| **History of prior TBI** | 79 (31) | | | >0.9 |
| > 1 | | 1 (1.8) | 3 (2.6) | |
| 1 | | 9 (16) | 17 (15) | |
| None | | 47 (82) | 97 (83) | |
| **Number of comorbidities, mean (SD)** | 3 (1.2) | 5 (3) | 5 (3) | >0.9 |
| **Number of medications, mean (SD)** | 0 (0) | 14 (9) | 15 (11) | 0.8 |
| **Anticoagulants** | 7 (2.8) | 24 (25) | 31 (21) | 0.4 |
| **Platelet aggregation inhibitors** | 15 (5.9) | 28 (30) | 44 (31) | 0.9 |
| **Pre-injury cognition** | 58 (23) | | | **<0.001** |
| Unimpaired | | 33 (60) | 66 (47) | |
| Mild cognitive impairment | | 8 (15) | 60 (43) | |
| Dementia | | 14 (25) | 14 (10) | |
| **ADL (Activities of Daily Living, range 1-27 (worst)), mean (SD)** | 22 (8.7) | 3 (5) | 4 (6) | 0.8 |
| **GFI (Groningen Frailty Indicator, range 0-15 (worst)), mean (SD)** | 45 (18) | 4 (3) | 4 (3) | 0.7 |
| **Injury mechanism** | 2 (0.8) | | | 0.5 |
| Ground level fall | | 58 (60) | 94 (61) | |
| Fall from height | | 14 (14) | 31 (20) | |
| Motor vehicle accident | | 21 (22) | 24 (16) | |
| Other | | 4 (4.1) | 5 (3.2) | |
| **Glasgow Coma Scale** | 0 (0) | | | 0.9 |
| 15 | | 72 (73) | 107 (69) | |
| 14 | | 23 (23) | 39 (25) | |
| 13 | | 4 (4.0) | 8 (5.2) | |
| **Loss of consciousness (LOC)** | 6 (2.4) | 42 (44) | 62 (41) | 0.7 |
| **Post-traumatic amnesia (PTA)** | 6 (2.4) | 45 (47) | 69 (46) | 0.9 |
| **Alteration of consciousness (AOC)** | 8 (3.2) | 40 (43) | 57 (38) | 0.5 |
| **TBI symptoms (LOC/PTA/AOC)** | 8 (3.2) | 60 (64) | 94 (62) | 0.8 |
| **Extracranial injury** | 3 (1.2) | 39 (40) | 77 (51) | 0.093 |
| **Injury Severity Score (ISS)** | 51 (20) | 10 (7) | 14 (12) | **0.025** |
| **CT positive for intracranial trauma** | 0 (0) | 50 (51) | 95 (62) | 0.079 |
| **ED hemoglobin (mg/dL)** | 12 (4.7) | 13.18 (1.69) | 13.16 (1.85) | >0.9 |
| **ED glucose (mg/dL)** | 13 (5.1) | 132 (69) | 130 (45) | 0.2 |
| **Hypotension (SBP < 90 mmHg)** | 0 (0) | 10 (10) | 8 (5.2) | 0.14 |
| **Hypoxia (SpO2 < 90%)** | 0 (0) | 7 (7.1) | 9 (5.8) | 0.7 |
| **ED disposition** | 0 (0) | | | 0.9 |
| ED Discharge | | 19 (19) | 33 (21) | |
| Hospital admit no ICU | | 51 (52) | 75 (49) | |
| Hospital admit with ICU | | 29 (29) | 46 (30) | |

| | | | | |
|---|---|---|---|---|
| **Discharge disposition for survivors** | 18 (7.1) | | | 0.4 |
| Home | | 69 (71) | 104 (75) | |
| Other hospital/SNF/Rehab unit | | 25 (26) | 33 (24) | |
| LTAC/Nursing home | | 3 (3.1) | 1 (0.7) | |
| **Study partner age, mean (SD), y** | 82 (32) | 62 (14) | 65 (12) | 0.2 |
| **Study partner Caucasian** | 53 (21) | 39 (56) | 98 (75) | **0.004** |
| **Study partner education, mean (SD), y** | 90 (36) | 15.49 (2.67) | 15.61 (2.59) | 0.7 |
| **Number of years study partner knows participant, mean (SD)** | 86 (34) | 43 (20) | 46 (16) | 0.6 |
| **Study partner's relationship with the participant** | 42 (17) | | | 0.062 |
| Spouse | | 30 (43) | 71 (50) | |
| Adult child | | 31 (44) | 40 (28) | |
| Friend/relative/sibling/neighbor | | 8 (11) | 29 (21) | |
| Paid caregiver/healthcare provider | | 1 (1.4) | 1 (0.7) | |
| **Study partner frequency of contact with the participant** | 78 (31) | | | 0.7 |
| Daily | | 47 (82) | 88 (75) | |
| 3 times per week | | 5 (8.8) | 11 (9.3) | |
| Weekly | | 5 (8.8) | 18 (15) | |
| Less than once a month | | 0 (0) | 1 (0.8) | |
| **Study partner reported caregiver strain at baseline** | 115 (45) | 16 (40) | 45 (46) | 0.5 |
| **Study partner reported caregiver strain during the study** | 110 (43) | 22 (55) | 69 (67) | 0.2 |

[1]n (); Mean (SD)

[2]Pearson's Chi-squared test; Wilcoxon rank sum test; Fisher's exact test

ED, Emergency Department; ICU, Intensive care unit; LTAC, Long-Term Acute Care facility; SBP, Systolic Blood Pressure; SNF, Specialized Nursing Facility; SPO2, blood Oxygen Saturation; TBI, Traumatic Brain Injury; y, years.

**eTable 2: Reliable change from 2-weeks to 6-months among older adults with TBI who survived until 2-weeks**

| | No. (%) | | | | | | |
|---|---|---|---|---|---|---|---|
| | | Age group | | | Pre-injury cognition | | |
| | **Overall** N = 179[1] | **Young-old (65-74)** N = 75[1] | **Middle-old (75-84)** N = 68[1] | **Oldest-old (85+)** N = 36[1] | **Unimpaired** N = 83[1] | **MCI** N = 59[1] | **Dementia** N = 16[1] |
| **GOSE reliable change** | | | | | | | |
| Improvement | 61 (46) | 29 (53) | 24 (46) | 8 (32) | 31 (51) | 25 (46) | 3 (30) |
| None | 58 (44) | 24 (44) | 25 (48) | 9 (36) | 27 (44) | 22 (41) | 4 (40) |
| Deterioration | 13 (9.8) | 2 (3.6) | 3 (5.8) | 8 (32) | 3 (4.9) | 7 (13) | 3 (30) |
| **GOSE change** | | | | | | | |
| Median (IQR) | 0 (0, 2) | 1 (0, 2) | 0 (0, 2) | 0 (-2, 1) | 1 (0, 1) | 0 (-1, 2) | 0 (-2, 1) |
| Range | -5, 5 | -4, 4 | -2, 5 | -5, 4 | -2, 5 | -5, 5 | -4, 1 |
| **ADL reliable change** | | | | | | | |
| Improvement | 44 (40) | 20 (40) | 19 (43) | 5 (29) | 21 (42) | 18 (39) | 0 (0) |
| None | 61 (55) | 28 (56) | 22 (50) | 11 (65) | 28 (56) | 25 (54) | 5 (71) |
| Deterioration | 6 (5.4) | 2 (4.0) | 3 (6.8) | 1 (5.9) | 1 (2.0) | 3 (6.5) | 2 (29) |
| **ADL change** | | | | | | | |
| Median (IQR) | -1 (-5, 0) | -2 (-4, 0) | -1 (-8, 1) | -1 (-6, 0) | -1 (-7, 0) | -2 (-4, 1) | 0 (-1, 6) |
| Range | -21, 11 | -19, 6 | -21, 11 | -17, 5 | -21, 5 | -16, 10 | -1, 11 |
| **Walking blocks reliable change** | | | | | | | |
| Improvement | 12 (11) | 5 (10) | 6 (13) | 1 (5.3) | 9 (17) | 3 (6.7) | 0 (0) |
| None | 68 (60) | 27 (54) | 26 (58) | 15 (79) | 26 (49) | 32 (71) | 6 (75) |
| Deterioration | 34 (30) | 18 (36) | 13 (29) | 3 (16) | 18 (34) | 10 (22) | 2 (25) |
| **Walking blocks change** | | | | | | | |
| Median (IQR) | 0 (0, 6) | 1 (0, 6) | 0 (0, 6) | 0 (0, 2) | 0 (0, 6) | 0 (0, 3) | 0 (0, 4) |
| Range | -24, 60 | -12, 60 | -21, 58 | -24, 7 | -24, 36 | -21, 60 | 0, 11 |
| **FAQ reliable change** | | | | | | | |
| Improvement | 19 (18) | 6 (13) | 8 (20) | 5 (31) | 6 (13) | 9 (21) | 2 (25) |
| None | 79 (76) | 39 (83) | 30 (73) | 10 (63) | 39 (85) | 30 (70) | 5 (63) |
| Deterioration | 6 (5.8) | 2 (4.3) | 3 (7.3) | 1 (6.3) | 1 (2.2) | 4 (9.3) | 1 (13) |
| **FAQ change** | | | | | | | |
| Median (IQR) | 0 (-3, 1) | 0 (-3, 0) | 0 (-2, 1) | 0 (-7, 1) | 0 (-1, 1) | -1 (-4, 0) | 1 (-4, 3) |
| Range | -30, 13 | -20, 6 | -19, 13 | -30, 5 | -20, 5 | -19, 10 | -30, 13 |
| **CDR sum of boxes + language and** | | | | | | | |

| | | | | | | | |
|---|---|---|---|---|---|---|---|
| **behavior reliable change** | | | | | | | |
| Improvement | 0 (0) | 0 (0) | 0 (0) | 0 (0) | 0 (0) | 0 (0) | 0 (0) |
| None | 65 (88) | 32 (91) | 24 (86) | 9 (82) | 32 (91) | 30 (88) | 3 (60) |
| Deterioration | 9 (12) | 3 (8.6) | 4 (14) | 2 (18) | 3 (8.6) | 4 (12) | 2 (40) |
| **CDR sum of boxes + language and behavior change** | | | | | | | |
| Median (IQR) | 0.0 (0.0, 1.0) | 0.0 (0.0, 0.5) | 0.5 (-0.5, 2.0) | 0.0 (0.0, 2.5) | 0.0 (0.0, 0.5) | 0.0 (-0.5, 1.5) | 2.5 (0.5, 3.5) |
| Range | -2.5, 13.5 | -1.0, 9.0 | -2.5, 13.5 | -1.0, 7.5 | -1.0, 7.5 | -1.0, 12.5 | -2.5, 13.5 |

[1]n ()

Abbreviations: ADL, Activities of daily Living; CDR, Clinical dementia rating; FAQ, Functional Activities Questionnaire; GOSE, Glasgow Outcome Scale Extended; MCI, mild cognitive impairment.

Sample size used per outcome (n): GOSE (132), ADL (111), Walking blocks (114), FAQ (104), CDR sum of boxes + language and behavior (74).
